# Trajectories of depressive symptoms and associated factors among reproductive age women in Khwisero, Western Kenya: a longitudinal community-based study

**DOI:** 10.1101/2025.11.05.25339562

**Authors:** Amanuel Abajobir, Estelle Sidze, Wendy Janssens, Nursena Aksünger, Daniel Maina, Mulusew J. Gerbaba

## Abstract

**Background:** In sub-Saharan Africa (SSA), maternal depression remains a public and social challenge. Available studies from high-income countries suggest that affected women do not constitute one homogeneous group in terms of severity, chronicity, and onset of symptoms. However, few studies have captured the distinct characteristics of such depression patterns in SSA. This study adds to the literature by examining depressive symptoms trajectories and associated factors among reproductive age women in Khwisero, western Kenya. It provides a unique contribution by generating evidence from community-based longitudinal data, using a semiparametric, group-based trajectory modeling to identify heterogenous subgroups of women, each following a distinct pattern of depressive symptom development over time, with their own trajectory pattern and growth parameters.

**Methods:** We used data collected from community-based longitudinal household surveys from 2019 to 2023. A total of 257 reproductive age (18–49 years) women were assessed for depressive symptoms using Center for Epidemiological Studies Depression (CES-D) scores and included in the analyses. Latent class growth curve mixture and generalized estimating equation models were used to identify trajectories of depressive symptoms and examine factors associated with each trajectory group, respectively. Bayesian information criterion, the probability of group memberships, average posterior probability, odds of correct classification, and biological plausibility were used to identify and determine the trajectory groups. A p-value of 0.05 was set to examine associations of each trajectory with factors.

**Results:** We found four distinct patterns of depressive symptoms among reproductive age women, namely moderately stable depression (19.8%, Group 1), mildly stable depression (56.8%, Group 2), higher but improving depression (18.6%, Group 3), and unstably high depression (4.8%, Group 4). We also found that reproductive age women who reported better self-rated health status and food security were less likely to experience unstable or moderately increasing depressive symptoms.

**Conclusion:** Reproductive age women exhibit distinct forms of depressive symptoms trajectories, underscoring the need to recognize and address heterogeneity in design, development, and delivery of maternal mental health interventions. These findings also provide deeper insights into the dynamic nature of depressive symptoms in under-researched rural African settings. Further research should focus on examining the effect of time varying factors and understanding potential mechanisms of underlying, proximal, and early life factors, including time varying factors, in a more diversified population using life course model and/or socioecological model. The differential impact of belonging in distinct groups of trajectories on women’s future health and their children’s health, development, and nutritional outcomes also requires further investigations.

**Article Highlights:**

- This study identified four distinct trajectories of depressive symptoms among reproductive age women–moderately stable, mildly stable, initially high but improving, and persistently unstable at high levels.
- Women with more favorable self-rated health and greater food security demonstrated a lower likelihood of experiencing persistent or escalating depressive symptoms, emphasizing the impact of social and economic determinants on mental wellbeing.
- The presence of multiple depressive symptom trajectories highlights the necessity of personalized intervention strategies that acknowledge individual variations in mental health progression and risk exposure.
- Further investigation is required to examine the roles of dynamic, time-dependent influences, and early-life exposures on depressive trajectories using integrative life course and socioecological frameworks.
- A deeper understanding of how different depressive trajectories shape long-term maternal health and child development, and nutritional outcomes is essential for developing evidence-based public health policies and interventions.

## Background

Common maternal mental health problems are one of the public health challenges affecting a substantial number of women globally [1, 2], with maternal depression or depression symptoms refers to depressive symptoms occurring during pregnancy, the postpartum period, or among women of reproductive age more broadly. The magnitude of these problems ranges from 7% to 30% [3, 4]. This magnitude is much higher in low- and middle-income countries (LMICs). A recent meta-analysis reported that 16% and 20% of women suffer from depression during antenatal and postnatal periods, respectively [5]. Existing studies on perinatal depression in sub-Saharan Africa (SSA) highlight high prevalence of antepartum (26%) and postpartum (17%) [6]. Maternal depression is associated with a lower access to, and use of healthcare services [7], increased risk of preterm birth, low birth weight, poorer growth and child development [8, 9], poor emotional and behavioral problems [10], and neonatal/infant mortality, as well as preeclampsia, cardiovascular diseases, and cognitive decline among women [11] in both LMICs and higher-income countries. The consequences of maternal depression include behavioral, cognitive, and physical health problems among children [12, 13] to obstetric complications including preterm birth, low birth weight [14], and impaired mother-child relationships [13]. It is also associated with increased risk of cognitive delays, socioemotional and behavioral difficulties in infants, pre-school, and older children [15].

However, selecting proven and effective interventions necessitates understanding the variations in mental health-related symptoms across different groups, identifying when women are most at risk, and determining the factors associated with the onset, severity, and chronicity of the disorder [16]. There is growing evidence that maternal depressive symptoms are heterogeneous, highly diversified with their onset, course, duration, and severity [17–19]. A recent systematic review of perinatal depressive symptom trajectories found distinct patterns of depressive trajectories across studies and emphasized the need for further research within different settings [14]. For example, while postnatal depression may resolve over time for some women, it may persist if left untreated in others well into their children’s preschool years [20]. Previous studies examined the trajectories of maternal depressive symptoms over the period between late pregnancy up to 1 to 2/5 years postpartum [19, 23] and checked population trends, mental health stability, and its predictors [19, 25].

Although depressive symptoms vary across individuals over time, most studies have failed to account for this variation [1] and there remain gaps in understanding what determines maternal depression trajectories in SSA. This may be due to the fact that most research on the trajectories of maternal mental health symptoms and determinants is based on cross-sectional studies, population average models, mixed effects models, and conventional growth models [6, 24]. Moreover, there are several limitations in the knowledge base on this subject. First, previous studies explored the rate of mental health change using “averaged means” such as longitudinal mixed-effects and latent growth curve models to examine individual variabilities of these symptoms over time using a single growth curve. This approach assumes that all women within a population follow the same pattern of growth curve over time [26]. However, existing evidence suggests heterogeneity in time of onset and progression of depressive symptoms. Therefore, previous findings might mask group differences and fail to quantify individual heterogeneity in mental health trajectory, including time of onset, and progression of depressive symptoms. Second, the literature did not examine the degree of individual fluctuations in mental health status or inform about the age at which women might be identified as being on different trajectories. Thirdly, such methods could not identify risk factors peculiar to each mental health trajectory. In another word, the risks of exposure might differ across groups which may affect their trajectory across their life course. Whereas the predictors for maternal depression are well documented, there remains a paucity of research that links longitudinal trajectories of depressive symptoms in reproductive age women to their risk factors. This is particularly important as some risk factors may be associated with certain subgroups of women with depressive symptoms, which would allow for targeted interventions [21]. Thus, it is essential to explore the trajectories of maternal depressive symptoms not only to recognize which women are targeted by early mental health interventions but also to identify modifiable risk factors. On the other hand, given the far-reaching consequences of depression on women and their children/families, a greater understanding of the course of symptoms over time is warranted. Therefore, using alternative group-based trajectory modeling, the primary objective of this study is to identify longitudinal patterns of depressive symptom trajectories and examine how key determinants are associated with trajectory group membership among reproductive age women in rural Kenya. These determinants may influence the onset, persistence, or resolution of depressive symptoms over time. This study uniquely contributes to the literature by generating evidence from community-based longitudinal data and by applying a semiparametric, group-based trajectory modeling approach that enables the identification of heterogeneous subgroups of women, each following a distinct depressive symptoms trajectory over time with unique shapes and growth parameters. This study has several policy implications. First, it provides new information on mental health status during women’s life course using community-based, panel data with repeated measurements in Kenya, a country experiencing demographic and epidemiologic transitions across time. This is particularly important in the case of maternal depression as it allows for capturing the diversity of these symptoms in terms of onset, course, timing, and severity [27]. Understanding such phenomena is important to determine interventions for preventing risks of mental health conditions that would occur later in life or to tackle the long-term consequences of mental health problems on maternal and child wellbeing. Second, it helps in identifying and targeting a group of women with distinct but similar characteristics of mental health dynamics and explores the differential role of social determinants of health in shaping the trajectory.

## Methods

### Study design and sampling

This analysis draws data from a community-based longitudinal survey conducted as part of the evaluation of the Innovative Partnership for Universal Sustainable Healthcare (*i*-PUSH) program among women from socioeconomically disadvantaged rural households in Khwisero subcounty, Kenya. The protocol for this “parent” study was registered (AEA Registry [AEARCTR-0006089] and ClinicalTrials.gov [NCT04068571]). Three additional waves of data were collected as part of an embedded study funded by the European and Developing Countries Clinical Trial program (EDCTP) and registered with the Pan African Clinical Trails Registry (PACTR202204635504887).

The longitudinal study began on 18/11/2019 and ended on 31/08/2024, by enrolling a cohort of rural low-income households which has reproductive age women, with under 4 years old children at recruitment. The sample size for the entire project was calculated based on the expectation that the *i*-PUSH program would produce a meaningful effect size of 0.4 in terms of healthcare utilization, with an intracluster correlation (ICC) of 0.014 [29]. A total of 24 villages were randomly selected from lists of 239 villages around two different health facilities. After conducting a census in each selected village to identify all the households meeting the selection criteria, 10 households were randomly selected from each village. In each household, adults, including the target women, reported on various health-related events, including depressive symptoms and healthcare service utilization. For the current analysis, we used data from a subgroup consisting of 257 women who participated in the endline survey (August 2024), had children aged 5 years or younger, and had consistent data across the entire follow-up period. The details of sample size computation and sampling procedures were described elsewhere in previous studies [28, 29]. Given the small sample sizes within specific subgroups (e.g., pregnant or early postpartum) and the dynamic nature of participants’ status across follow-up (e.g., transitioning from pregnancy to postpartum), we retained a broader categorization of ‘reproductive age women’ to preserve statistical power and ensure stable trajectory modeling. All study participants, irrespective of intervention status, received appropriate psychosocial support, as deemed ethically necessary, throughout the data collection and follow-up period.

### Data collection

#### Outcome variables

A reproductive woman age (18–49 years) who had at least one under-five-year-old child was invited to respond to our questions related to a mental health questionnaire. A total of 16 waves of mental health data were collected using self-reported 10-item Center for Epidemiological Studies Depression (CES-D) scale [30, 31]. Ten items on four scales were used to ask participants to rate how often they had conflicting feelings over the course of the past seven days on four scales: (1) never; (2) a little of the time (1–2 days); (3) a moderate amount of the time (3–4 days); and (4) most or all of the time (5–7 days). Our study sample included women across a range of life stages–pregnant, postpartum, and non-pregnant–throughout the study follow-up period, with a limited sample size within these subgroups. As such, we did not disaggregate the sample by pregnancy status and consistently used ‘depressive symptoms among women of reproductive age group’ consistently to best capture the scope and intent of the analysis.

#### Explanatory variables

We also measured contextual factors, such as family/household socioeconomic status, wealth, availability of formal and informal supports, health literacy, mother’s age (in years), parity, literacy, education, income, marital status, the presence of a father in the home, female-headed household, residence, employment, parental education, poor physical health, and behavioral factors. The wealth index of households was calculated using information on the ownership of certain assets by households. Additionally, the analyses considered mothers’ history of chronic illnesses and decision-making power. Mothers’ decision-making power was assessed through a binary (yes/no) question asking who determined whether the mother or child should seek medical care when ill. The details of the methods used to assess these risk factors have been described in previous studies [28, 29].

#### Data quality control

There were strict data quality control measures in place, such as pretest to fix any issues that arose with the tools, the participants, or the field environment. Real-time data collection in the field was supervised by trained team leaders, who also ensured data quality by conducting frequent spot checks and sit-ins on up to 5% of each field worker’s daily work. By editing the data before they were recorded, they also validated the accuracy of the data collected before sending it to the database. Additionally, an automated tool verified the accuracy, consistency, and completeness of the data.

#### Data analyses

The analysis was based on 257 women of reproductive age–representing 73% of the total original sample–who had at least three-time points of measurement for reliable trajectory modeling. Latent class growth curve mixture and multinomial logistic regression models were used to identify distinct trajectories of depressive symptoms and explore factors associated with depressive symptoms, respectively. The predictors of depressive symptoms trajectories were categorized into sociodemographic and psychosocial factors. First, exploratory data analysis of depressive symptoms trajectory and individual factors plots were graphed to examine the shape and identify potential outliers, respectively. Descriptive statistics of explanatory variables are presented using mean and standard deviations for continuous variables and percentage for categorical variables. We identified subgroups of distinct depressive symptom trajectories using latent class growth mixture model (LCGMM) [32] to capture the heterogeneity in depressive symptoms development over time in k (4) number of subgroups (classes) each with distinct mental health trajectory shape and their own growth parameters (i.e., cubic, quadratic, linear slope, and intercept). A LCGMM is a semiparametric mixture, group-based modeling approach used to identify the different trajectories. It is a form of finite mixture modeling where the method is designed to identify groups/clusters of individuals following similar developmental trajectories. The models were estimated using the Stata “traj” command without a priori hypothesis regarding the existence of distinct trajectory and respective number and shape to flexibly handle unbalanced longitudinal data and allow model fit assessment in a user-friendly manner. We selected the final model after running several models starting from linear function, followed by addition of quadratic and cubic slopes (to allow the curved model developmental patterns), respectively. To determine the optimal number of trajectories, a single class trajectory model was fitted assuming all individuals follow a similar trajectory over time. Then, subsequent models were run by adding one additional trajectory group (classes) at a time to investigate whether the model fitness improves due to additional class(es) or not. The final number of trajectory groups were determined based on model fit indices such as Bayesian information criterion (BIC), Bayes factor, average posterior probabilities, the odds of correct classification (OCC), the proportion of group membership, shapes of the trajectories, and the biological plausibility [32]. As such, we selected the four-trajectory group due to its better statistical fit and enhanced interpretability, although fit indices and classification diagnostics showed only marginal differences between the models. After identifying the optimal number of trajectories, the shapes of individual trajectory were determined. Initially, the cubic function was used for all trajectory groups. Then, in each group, step-by-step lower order polynomial functions (quadratic followed by linear and constant functions) were considered until BIC indicated improved fit and we reached a function containing all growth parameter estimates with a significant p-value (p < 0.05). Finally, the association between depressive symptoms group and associated factors was assessed using a multinomial logistic regression model. We used a “mild and stable depressive symptoms status” as a reference category. Therefore, the regression estimates can be interpreted as the ratio of the probability of choosing one outcome category over the reference outcome. In the two-step model, first the association between depressive symptom trajectories and each determinant was examined (Model I). Subsequently, multivariate analysis was conducted (Model II). To assess the robustness of the model, multivariable generalized estimating equation (GEE) models were employed to predict higher depressive symptoms scores by utilizing a Gaussian family, identity link, exchangeable correlation structure, and robust standard errors, clustered by participant. Although model selection was informed by these fit indices and biological plausibility, some inconsistencies may still remain, which may have led to over- or underestimation of trajectory numbers and group membership.

#### Compliance with ethical standards

This study involved human participants and was conducted in accordance with the Declaration of Helsinki. Participants were given a written informed consent form, gave their consent, and signed before the interview and after enumerator described the objectives of the study, key project details, potential risks and benefits for participants, expectations for privacy and confidentiality, and contact information and request their consent to participate. Each participant was free to choose whether to participate in the study, and they were also given the option to opt out. If they were unable to write, they were given a stamp and/or a witness to sign to confirm the consent. Each participant received an anonymous number or ID, and the information we obtained from them was kept private. Study protocol was reviewed and approved by Internal Review Board of the African Population and Health Research Center and Amref Health Africa’s Ethical and Scientific Review Committee (ESRC) (ref. P679-2019). The nested study that served as the foundation for this analysis was also reviewed and approved by the ESRC (ref. P1060/2021). A research permit was granted by Kenya’s National Commission for Science, Technology, and Innovation (NACOSTI). Otherwise, there is no conflict of interest to declare.

## Results

### Baseline characteristics

The study sample was representative of reproductive age women in the setting [28], with a mean baseline CES-D score of 28 (range: 8-45). Table 1 provides demographic and socioeconomic characteristics of the study sample. We followed 257 women of reproductive age from eligible households to collect data for this study. The average age of mothers included in the study was 30.6 years, with most falling within the 25-34 age range (50.6%). The average age at first sexual intercourse was 17.4 years. Most respondents (91%) reported ever using some form of birth control, while 65.7% were currently using birth control methods. Most participants belonged to the Luhya ethnic group (88.3%). Additionally, approximately 46% of the respondents reported being employed and more than half of the participants (55.5%) had worked for money in the past year. When asked to rate their health compared to a year ago, 47.9% felt their health remained the same, while 22.2% believed it had somewhat worsened. Household hunger was assessed, 36.1% indicated experiencing hunger at some point in the last 12 months (Table 1).

**Table 1.**
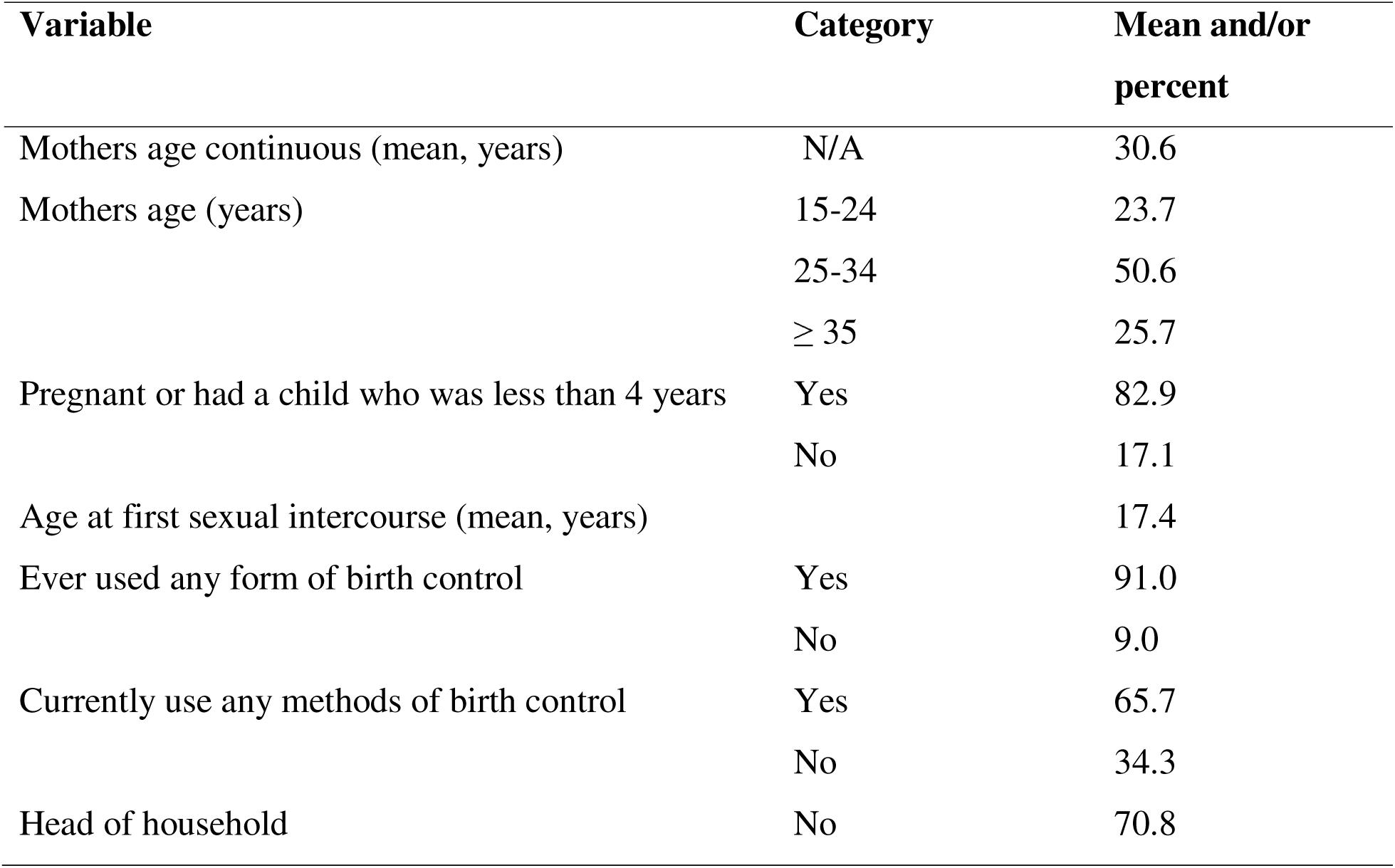
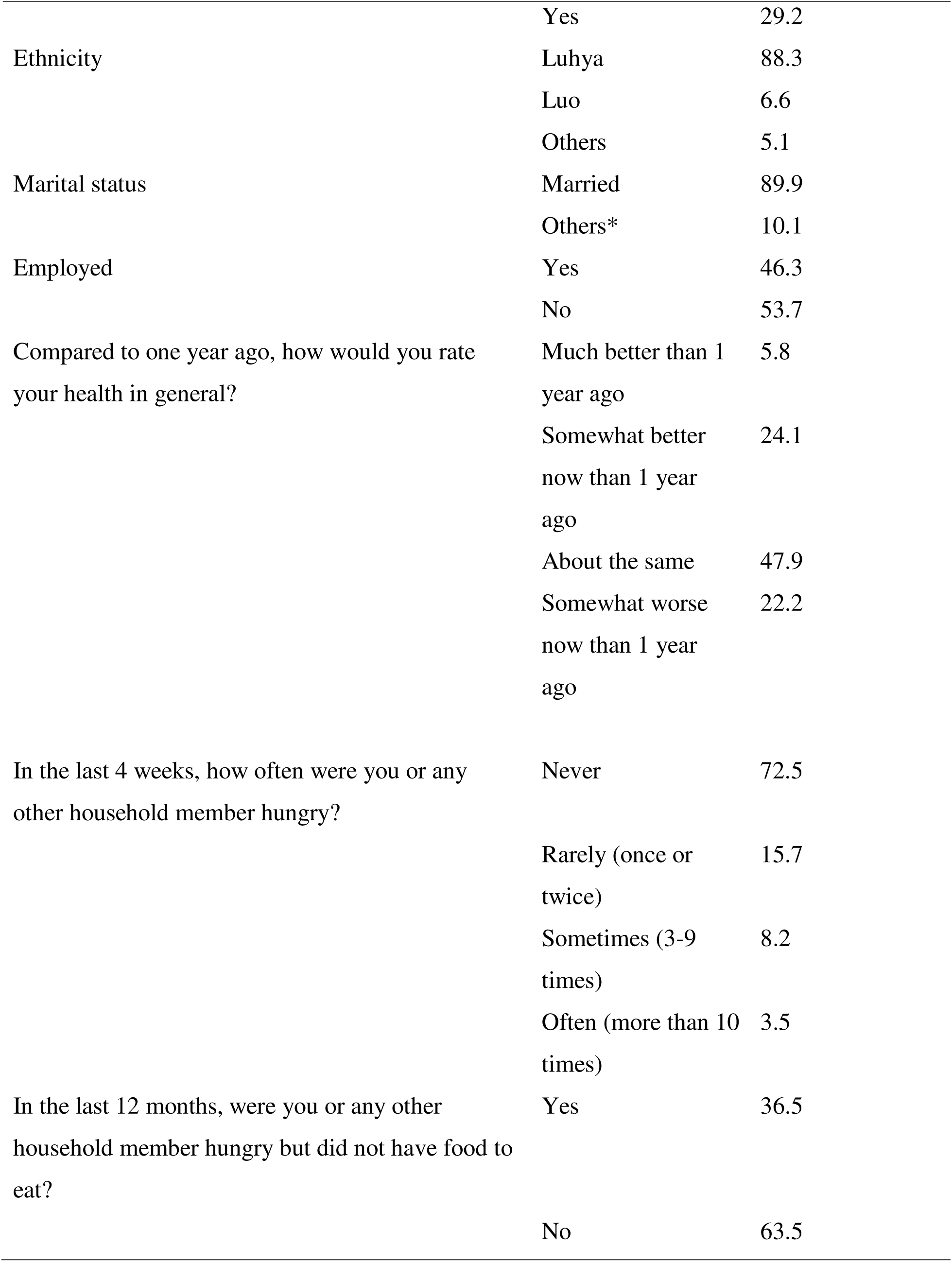
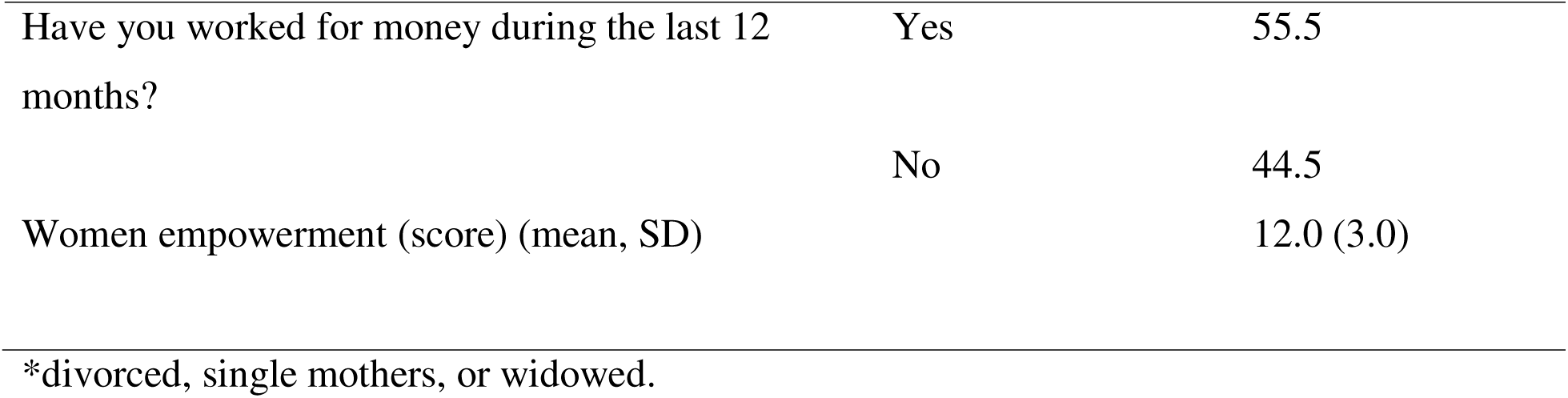
Baseline characteristics of study participants (N = 257)

### Identifications of depressive symptoms trajectories: model selection

Table 2 presents model fit indices of models we estimated to identify the number of classes of depressive symptoms trajectories that best describe depressive symptoms patterns of the population under study. Accordingly, the BIC continued to decrease as the number of classes increased. The average posterior probabilities (≥ 0.70), odds of correct classification (≥ 5), and the proportion of groups in each member were acceptable. Therefore, taking its biological plausibility/interpretability, and Bayes factors into account, we opted to identify 4 groups of depressive symptoms trajectories, which appeared to be optimal.

**Table 2.**
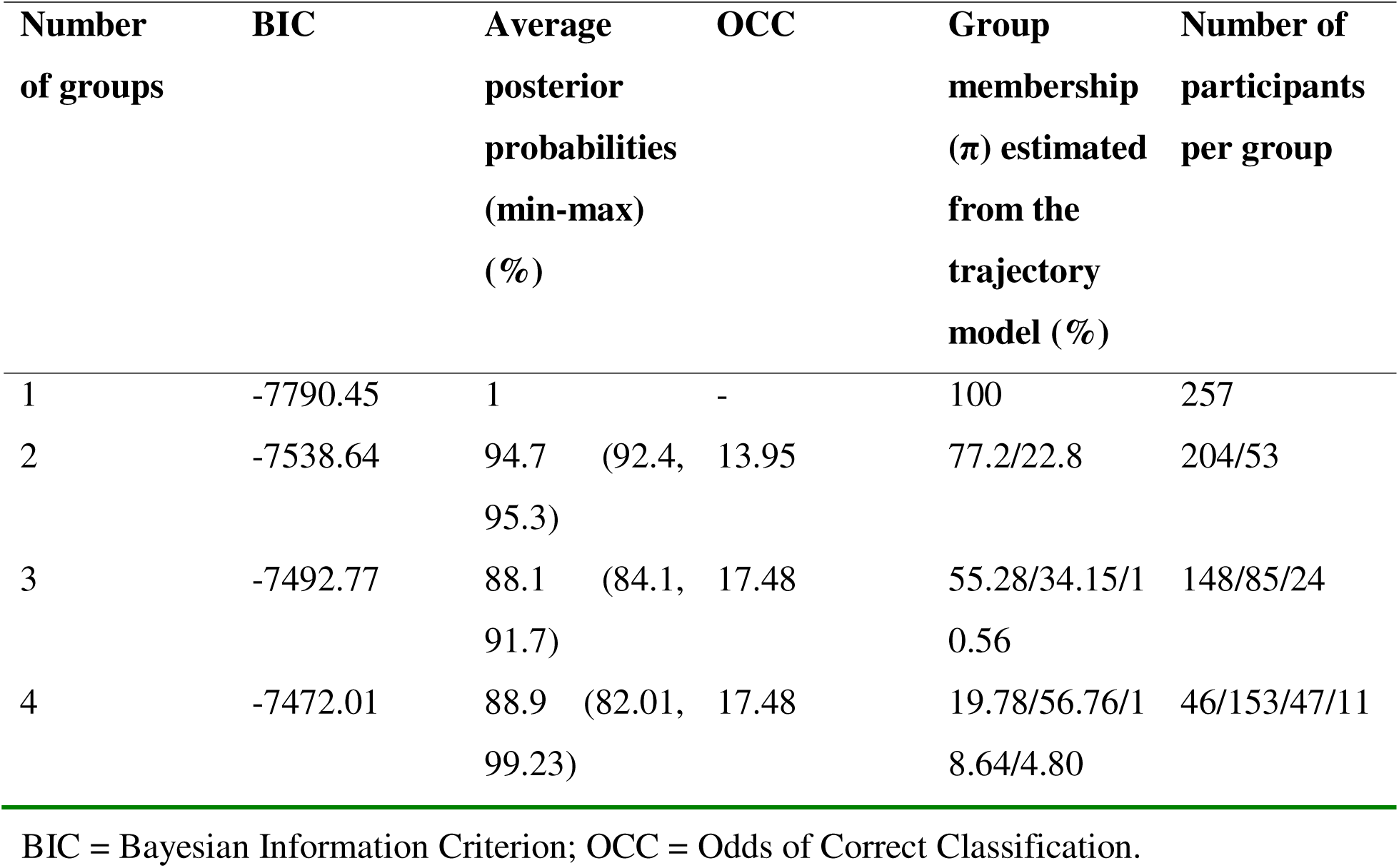
Model fit indices of group-based trajectory model.

### Shapes of depressive symptoms trajectories

Figure 1 shows the shape of depressive symptoms trajectories. Accordingly, there were groups with moderately stable depressive symptoms (19.8% Group 1); mildly stable depressive symptoms (56.8%, Group 2); higher but improving depressive (18.6% Group 3); and unstably high depressive symptoms (4.8%). As indicated in Figure 1, there were groups characterized by higher depressive symptoms scores between round 3 and 7, which might be associated with the first COVID-19 detection period where there was a systemic risk during that period. However, a small proportion of the study population had such unstable symptoms. The change in growth parameters of these symptoms were increasing through time, with a mean score of 0.0193 (SE: 0.005). In general, the result showed a complaint of at least 5 symptoms of depression by the study group (Table 3).

**Figure 1.**
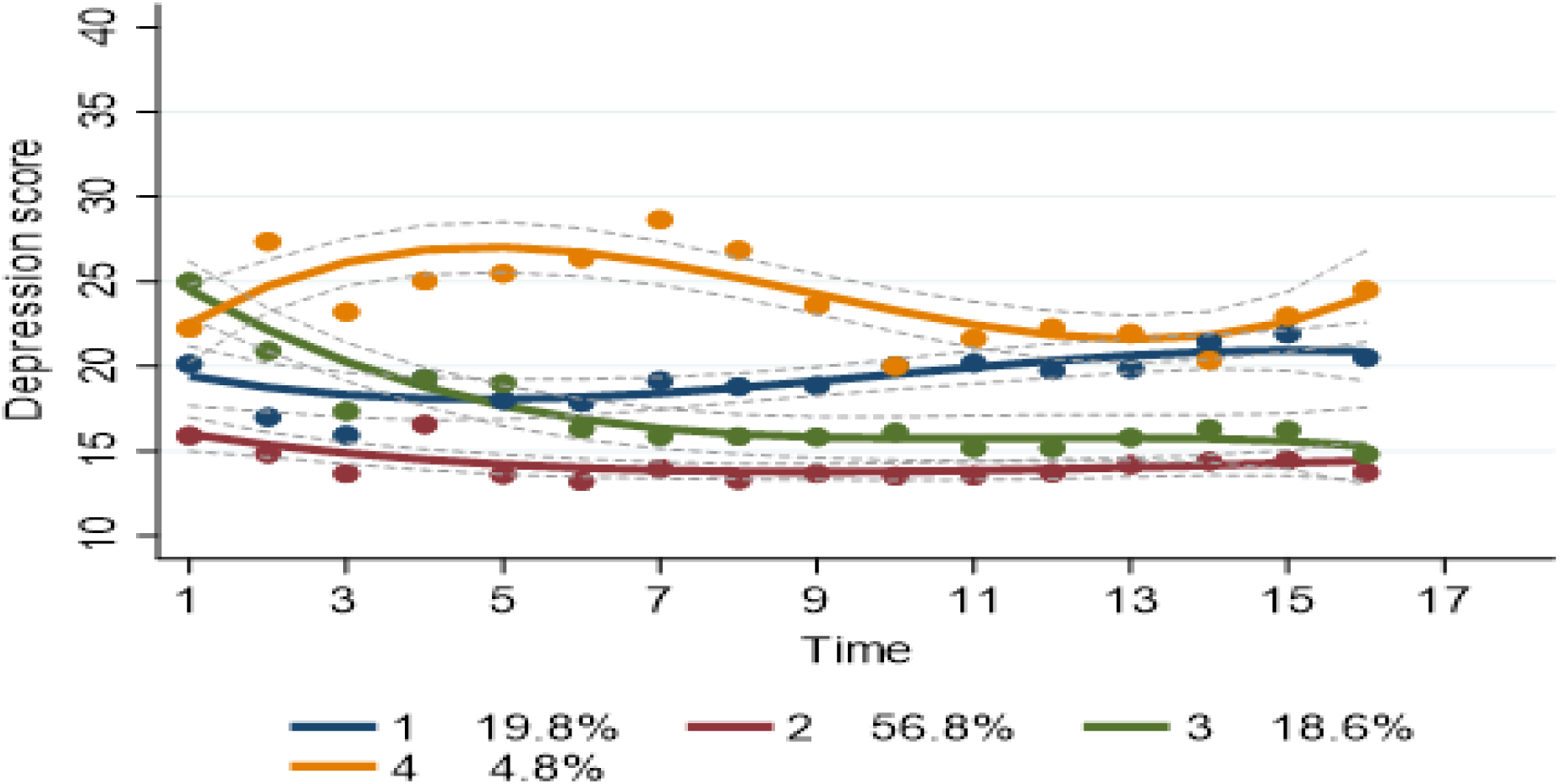
Depressive symptoms trajectories in reproductive age women in Khwisero, Kenya. Note: Group 1 is defined as mildly stable depressive symptoms; Group 2 as moderately stable depressive symptoms; Group 3 as moderately higher and unstable depressive symptoms; and Group 4 as relatively unstable higher depressive symptoms.

**Table 3.**
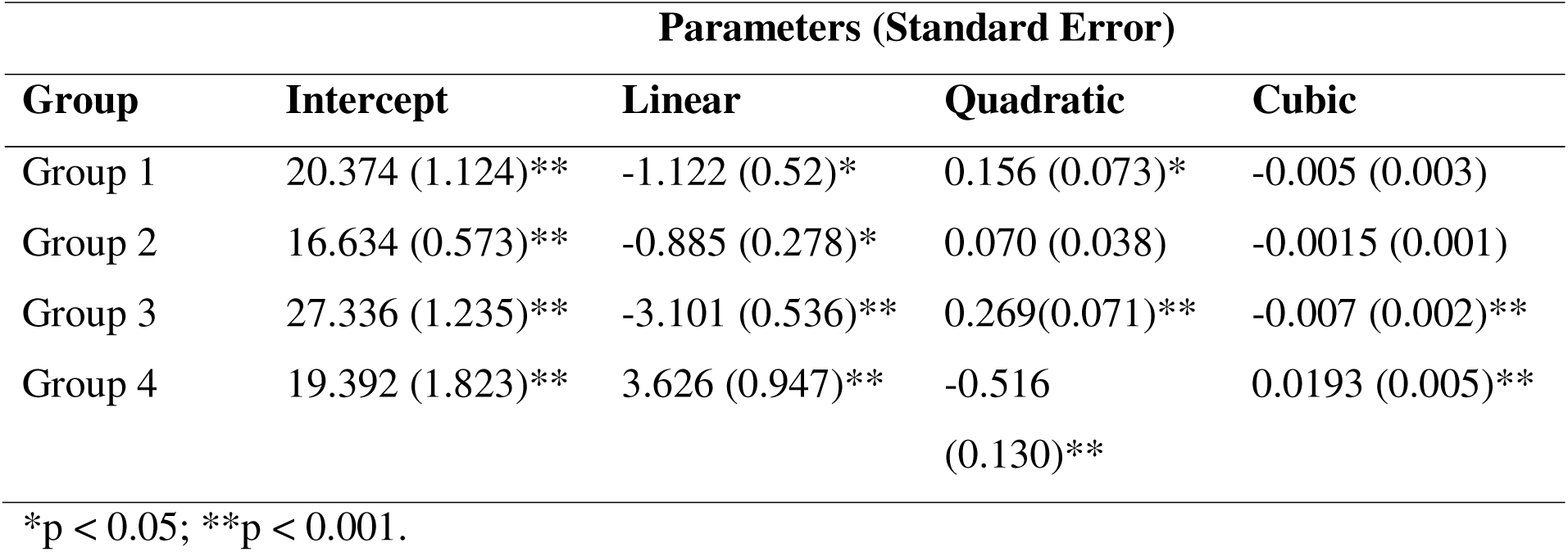
Growth parameter estimates of the selected model.

| Group | Parameters (Standard Error) |  |  |  |
| --- | --- | --- | --- | --- |
|  | Intercept | Linear | Quadratic | Cubic |
| Group 1 | 20.374 (1.124)** | -1.122 (0.52)* | 0.156 (0.073)* | -0.005 (0.003) |
| Group 2 | 16.634 (0.573)** | -0.885 (0.278)* | 0.070 (0.038) | -0.0015 (0.001) |
| Group 3 | 27.336 (1.235)** | -3.101 (0.536)** | 0.269(0.071)** | -0.007 (0.002)** |
| Group 4 | 19.392 (1.823)** | 3.626 (0.947)** | -0.516<br>(0.130)** | 0.0193 (0.005)** |

\*p < 0.05; \*\*p < 0.001.

### Risk factors associated with depressive symptoms trajectories

Table 4 summarizes baseline characteristics associated with trajectory membership relative to the low-stable depressive symptoms trajectory. First, we explored the independent association of each determinant with each group (Model I). In model I, household head status, food security, marital status, age, self-rated health, agriculture land ownership, and women empowerment were more likely to be associated with moderately increasing and an unstable depressive symptom compared to women who belonged to the stable depressive symptoms trajectory. No significant differences in depressive symptoms status were observed based on income, age, or educational status.

**Table 4.** Risk factors associated with trajectory memberships.

| **Group I** |  |  |  |  |
| --- | --- | --- | --- | --- |
| **Risk Factor** | **Univariate Model, OR (95% CI)** | **p-value** | **Multivariate Model, OR (95% CI)** | **p-value** |
| Maternal age | 1.012 (0.902-1.136) | 0.828 | 0.935 (0.678-1.290) | 0.683 |
| Food secure | 0.254 (0.040-1.609) | 0.146 | 0.237 (0.001-66.417) | 0.617 |
| Female-headed household | 2.389 (1.051-5.432) | 0.037 | 0.000 (0.000-0.001) | 0.942 |
| Marital status (married) | 0.092 (0.014-0.593) | 0.012 | 2.968 (0.958-9.197) | 0.998 |
| Own livestock | 2.983 (0.619-14.367) | 0.173 | 3.442 (0.042-285.385) | 0.583 |
| Employed (Yes) | 0.551 (0.255-1.190) | 0.129 | 0.285 (0.159-0.513) | 0.966 |
| Ethnicity | 0.257 (0.082-0.809) | 0.990 | 0.312 (0.157-0.620) | 0.997 |
| Women empowerment | 1.302 (0.814-2.084) | 0.272 | 1.088 (0.295-4.004) | 0.898 |
| Monthly household income | 1.000 (1.000-1.000) | 0.877 | 1.000 (1.000-1.000) | 0.579 |
| Own agriculture land | 1.083 (0.458-2.561) | 0.854 | 0.000 (0.000-0.000) | 0.949 |
| Attended school (secondary and above) | 0.589 (0.231-1.499) | 0.266 | 9.198 (0.031-2689.525) | 0.443 |
| Self-rated health | 0.552 (0.348-0.877) | 0.012 | 3.114 (0.110-87.868) | 0.504 |

| **Group III** |  |  |  |  |
| --- | --- | --- | --- | --- |
| **Risk Factor** | **Univariate Model, OR (95% CI)** | **p-value** | **Multivariate Model, OR (95% CI)** | **p-value** |
| Maternal age | 0.955 (0.885-1.031) | 0.110 | 0.997 (0.925-1.074) | 0.946 |
| Food secure | 0.336 (0.168-0.670) | 0.002 | 0.636 (0.200-2.017) | 0.439 |
| Female-headed household | 2.809 (1.205-6.552) | 0.017 | 0.662 (0.190-2.306) | 0.516 |
| Attended school (secondary and above) | 1.278 (0.540-3.021) | 0.576 | 1.251 (0.417-3.749) | 0.689 |
| Marital status (married) | 0.169 (0.047-0.605) | 0.006 | 0.490 (0.035-6.803) | 0.595 |
| Own livestock | 1.164 (0.546-2.481) | 0.693 | 0.563 (0.159-1.998) | 0.374 |
| Employed (Yes) | 0.954 (0.877-1.038) | 0.913 | 1.554 (0.490-4.930) | 0.455 |
| Ethnicity | 0.994 (0.977-1.012) | 0.491 | 0.448 (0.024-8.221) | 0.588 |
| Women empowerment | 1.119 (0.985-1.270) | 0.084 | 1.280 (0.947-1.731) | 0.108 |
| Monthly household income | 1.000 (0.994-1.006) | 0.135 | 1.000 (1.000-1.000) | 0.155 |
| Own agriculture land | 2.024 (0.876-4.674) | 0.099 | 2.570 (0.705-9.371) | 0.152 |
| Self-rated health | 0.479 (0.283-0.809) | 0.006 | 0.462 (0.246-0.866) | 0.016 |

| **Group IV** |  |  |  |  |
| --- | --- | --- | --- | --- |
| **Risk Factor** | **Univariate Model, OR (95% CI)** | **p-value** | **Multivariate Model, OR (95% CI)** | **p-value** |
| Maternal age | 1.047 (0.989-1.108) | 0.123 | 1.018 (0.923-1.123) | 0.709 |
| Food secure | 0.067 (0.014-0.322) | 0.008 | 0.274 (0.069-1.086) | 0.045 |
| Female-headed household | 0.000 (0.000-0.000) | 0.987 | 0.628 (0.123-3.192) | 0.574 |
| Marital status (married) | 1178791.124 (950173.615-1462415.385) | 0.990 | 0.859 (0.051-14.446) | 0.915 |
| Own livestock | 2.401 (0.700-8.239) | 0.164 | 2.151 (0.516-8.961) | 0.292 |
| Employed (Yes) | 2.044 (0.512-8.156) | 0.311 | 0.240 (0.038-1.505) | 0.127 |
| Ethnicity | 0.944 (0.575-1.549) | 0.871 | 1.003 (0.984-1.023) | 0.739 |
| Women empowerment | 1.017 (0.834-1.240) | 0.860 | 0.896 (0.684-1.174) | 0.424 |
| Monthly household income | 1.000 (1.000-1.000) | 0.415 | 1.000 (1.000-1.000) | 0.523 |
| Own agriculture land | 1.131 (0.281-4.557) | 0.862 | 1.020 (0.224-4.651) | 0.978 |
| Attended school (secondary and above) | 0.913 (0.224-3.730) | 0.898 | 1.682 (0.396-7.145) | 0.481 |
| Self-rated health | 0.436 (0.211-0.902) | 0.025 | 0.736 (0.332-1.630) | 0.449 |

**Note:** Group II (stable depressive symptoms trajectory) is a reference category.
Group I: adjusted for: maternal age, food security status, household head, marital status, school attendance, own livestock ownership, employment, ethnicity, women empowerment, monthly household income, agriculture land ownership, and self-rated health.
Group III: adjusted for: maternal age, food security status, household head, marital status, school attendance, own livestock ownership, employment, ethnicity, women empowerment, monthly household income, agriculture land ownership, and self-rated health.
Group IV: adjusted for: maternal age, food security status, household head, marital status, school attendance, own livestock ownership, employment, ethnicity, women empowerment, monthly household income, agriculture land ownership, and self-rated health.

However, the multivariate analysis (Model II) showed only food security and those women who had self-rated status had reduced form of unstable depressive symptoms status compared with the stable depressive symptoms trajectory group. Educated women were also more likely to belong in an increasing stable depression group than the stable depressive symptoms trajectory, whereas living with a female-headed household did not predict membership to any of the trajectories.

**Figure 2:**
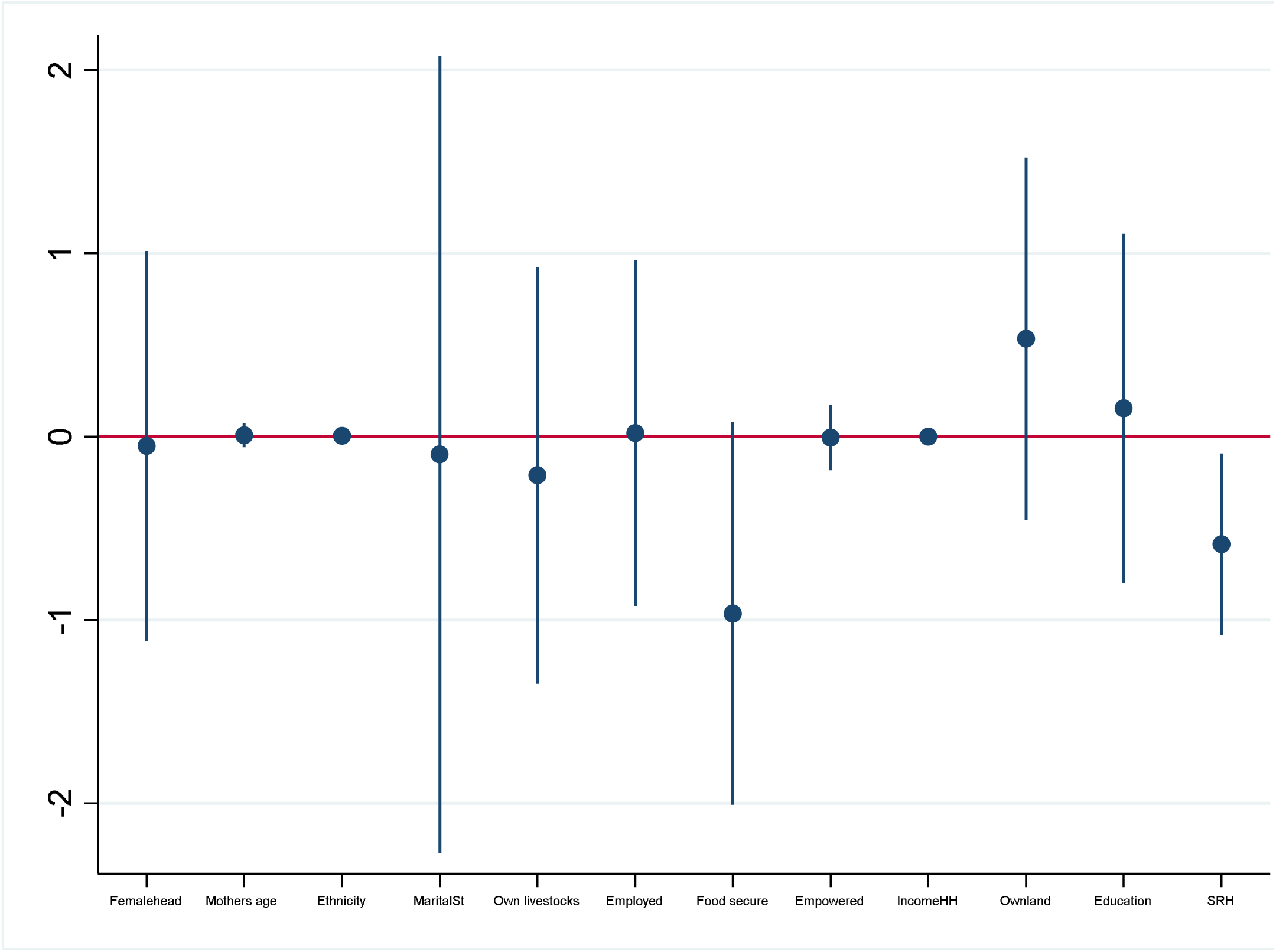
Association of factors with depressive symptoms trajectories.

## Discussion

Identifying groups of individuals following similar patterns of depressive symptoms is useful to understand the onset, pathways, and factors associated with the development of depression over time. This would help to understand the mechanisms underlying changes in depression status and develop tailored interventions based on distinct trajectories. To date, studies on depression trajectories are mainly from developed regions and available evidence from LMICs examining the risks of depression in reproductive age women is dependent on either single point of measurement or relied on conventional methods of longitudinal analysis by ignoring two main issues: dynamism of change in depression and the potential diversity/heterogeneity between individuals [33]. Therefore, we used LCGMM approach assuming women living in poor income settings could also exhibit a differential trajectory in depressive symptoms progression/development. To the best of the authors’ knowledge, this is the first study evaluating heterogeneity of depressive symptoms trajectories and its relationship with other social determinants of health using representative cohorts of reproductive age women living in rural Kenya.

In this study, we found differential numbers and shapes of depressive symptoms trajectories. Reproductive age women included in this study exhibited four multiple trajectories that fall within relatively mild to moderate depressive symptoms patterns over time. However, close to 5% of the women population exhibited unstable and higher depressive symptoms. Compared to previous studies in Kenya [33], we did not find severe forms of depressive symptoms over time, although comparison across studies is difficult due to the differences in study populations and developmental periods examined. Previous studies, including those from well-developed regions, usually report three to five trajectories mainly of chronic high depression, stable high depression, increasing depression, decreasing depression, and normal depression score. The differences might be due to the variations in contextual or environmental factors that may lead to differential in exposures and vulnerabilities among populations living in different regions. These differences in the trajectory patterns may also be influenced by life course and stress processes, highlighting how the timing and accumulation of stressors, alongside protective/resilient factors, shape mental health trajectories across different contexts [34]. In developed countries, for example, exposure to extreme conditions such as poverty have been associated with higher and unstable depression [35]. Studies from high income countries also showed that the highest depression started during adolescence coming from families with lower socioeconomic status [36]. We also estimated the impact of baseline characteristics on women’s depressive symptoms status and found women living in food secure environment and score better health status are less likely to experience depressive symptoms as compared to their counterparts.

This study possesses several strengths. We examined depressive symptoms changes during an important developmental period, reproductive age, in which important biological/physiological and environmental (e.g., food insecurity) changes occur. Second, we used a longitudinal representative sample of a cohort of reproductive age women who were born and have grown during a similar period in which important structural changes have occurred such as economic growth, volatile national food prices and food crises, climate change, etc. Thirdly, our analyses used an accelerated longitudinal design that allowed for assessing depressive symptoms changes over time spanning for three years among cohorts of reproductive age women with 16 rounds of a dataset. Such approaches are considered as a single cohort (i.e., it assumes no age by cohort e ects). Methodologically, the model was not imposed ex-ante but rather emerged from the data. To identify distinct patterns of depressive symptoms trajectories, we used data-driven person-centered growth mixture modeling approach to explore distinct patterns of depressive symptoms changes across periods. Such analysis illustrates the natural heterogeneity in the development of depression status through time. Finally, rather than heuristically categorizing depression status, we used continuous depressive symptoms score rather than dichotomous measures to avoid misclassification.

The findings of this study should be interpreted by taking account of important limitations. First, there was inconsistency in model fit, although we chose based on different model fit indices and its biological plausibility. Therefore, estimates of true membership and the number of trajectories among the studied populations could possibly be over or underestimated. Second, the final model was estimated using 73% of the sample were disadvantaged groups, it is, therefore, likely that excluded group may underestimate the relationship of the trajectories and the relationship of depressive symptoms with other social determinants of health, despite sensitivity analysis showing no differing patterns among the lost to follow-ups. The existence of trajectories we found in this study also warrants further validation using similar parametric or non-parametric spline-based methods that include investigation of the underlying mechanisms associated with development and maintenance of these trajectories. Additionally, we did not collect detailed data on pregnancy stage or exact child age, which limited our ability to capture qualitative differences in depressive symptoms and to disaggregate findings by maternal pregnancy status or child age. Furthermore, we did not collect data on genetic and environmental factors which would have explained the development of depression. We also acknowledge that we were unable to account for the contributions of time varying confounders or unmeasured factors on changes in depressive symptoms over time.

## Conclusions

This study identified distinct trajectories of depressive symptoms among reproductive age women. Given the trends of different contexts and a spectrum of structural changes in the country such as food price volatility, climate change, and COVID-19 pandemic that affect multitudes of life patterns, the findings highlight the presence of distinct trajectories and the possible persistence of depressive symptoms among certain groups of reproductive age women for extended period of time. Thus, any interventions designed in such a context should consider heterogeneity in depression development and emphasize the improvement of other social determinants of health such as food security status. These findings suggest the need for targeted mental health support within community and primary care systems to better address the mental health needs of at-risk women [37]. Further research should focus on examining the effect of time varying confounders and understanding potential mechanisms of underlying, proximal, early life factors and genetics in a more diversified population using life course model and/or socioecological model. The differential impact of belonging in distinct groups of trajectories on the future health, child health and development, and nutritional outcomes also requires further investigations.

## Authors’ contributions

AA designed the study and conceived the idea for this analysis; MJ designed and led the analysis and drafted the manuscript. All authors then revised critically for important intellectual content of the manuscript.

## Data Availability

Since they pertain to a “high-risk” population, the data are not generally accessible but will be provided upon request. Links to published full-text protocol, protocol registrations, and other resources are offered, nevertheless. The published protocol is available at: https://www.ncbi.nlm.nih.gov/pmc/articles/PMC8443110/.

## Acknowledgements

The research team is grateful to the funder, implementers, study participants, and field team, as well as county and subcounty officials and other stakeholders who contributed directly or indirectly to the study’s success.

